# Seroprevalence of SARS-CoV-2 Antibodies Among 925 Staff Members in an Urban Hospital Accepting COVID-19 Patients in Osaka Prefecture, Japan

**DOI:** 10.1101/2020.09.10.20191866

**Authors:** Tsuotmu Nishida, Hiromi Iwahashi, Kazuhiro Yamauchi, Noriko Kinoshita, Yukiyoshi Okauchi, Norihiro Suzuki, Masami Inada, Kinya Abe

## Abstract

**Background:** The subclinical severe acute respiratory syndrome coronavirus 2 (SARS-CoV-2) infection rate in hospitals during the pandemic remains unclear. To evaluate the effectiveness of our hospital’s current nosocomial infection control, we conducted a serological survey of the anti-SARS-CoV-2 antibody (immunoglobulin G) among the staff of our hospital, which is treating coronavirus disease 2019 (COVID-19) patients.

**Methods:** The study design was cross-sectional. We measured anti-SARS-CoV-2 immunoglobulin G in the participants using a laboratory-based quantitative test (Abbott immunoassay), which has a sensitivity and specificity of 100% and 99.6%, respectively. To investigate the factors associated with seropositivity, we also obtained some information from the participants with an anonymous questionnaire.

**Results:** We invited 1133 staff members in our hospital, and 925 (82%) participated. The mean age of the participants was 40.0±11.8 years, and most were women (80.0%). According to job title, there were 149 medical doctors or dentists (16.0%), 489 nurses (52.9%), 140 medical technologists (14.2%), 49 healthcare providers (5.3%), and 98 administrative staff (10.5%). The overall prevalence of seropositivity for anti-SARS-CoV-2 IgG was 0.43% (4/925), which was similar to the control seroprevalence of 0.54% (16/2970)) in the general population in Osaka during the same period according to a government survey conducted with the same assay. Seropositive rates did not significantly differ according to job title, exposure to suspected or confirmed COVID-19 patients, or any other investigated factors.

**Conclusion:** The subclinical SARS-CoV-2 infection rate in our hospital was not higher than that in the general population under our nosocomial infection control measures.

## Introduction

Coronavirus disease (COVID-19), which is caused by infection with severe acute respiratory syndrome coronavirus 2 (SARS-CoV-2), first appeared in Wuhan, China, in December 2019 and triggered a pandemic. Since COVID-19 emerged, an increasing number of people have contracted it and died around the world. Hospital staff are at the front line of the efforts to control the ongoing COVID-19 pandemic and are at high risk of infection with SARS-CoV-2, which is highly contagious.

Consequently, nosocomial SARS-CoV-2 infections in hospital staff can be problematic. Chu et al. reported that this disease is often diagnosed in medical staff who were not in charge of affected patients in a hot spot region of the pandemic ^1^.

As of July 14, 2020, a total of 12,964,809 people had been infected with SARS-CoV-2, and 570,288 people had died of COVID-19 worldwide, with a total of 22,220 confirmed cases and 980 deaths in Japan according to the World Health Organization (WHO) Situation Report –176 ^2^. Compared with the global situation, Japan has achieved relatively better control of the pandemic and has maintained a relatively low incidence of nosocomial infections in the hospital. However, there are few available data on the rate of seropositivity for SARS-CoV-2 antibodies in hospital staff in Japan. Our hospital is a medium-volume hospital with 613 beds, including 14 in the Infectious Disease Unit. It is in an urban area of Osaka Prefecture, Japan, and is a designated medical institution for type II infectious diseases. There are 351 medical institutions with 1,758 beds in Japan. At the same time, as one of the region’s essential hospitals, our hospital plays a significant role in community care. At the government’s request, we were the hospital to accept four asymptomatic COVID-19 patients from the cruise ship, the Diamond Princess, on February 22, 2020. After that, COVID-19 spread in Japan, and the number of patients admitted to our hospital gradually increased. Due to the pandemic, Osaka Prefecture requested an increase in the availability of beds for COVID-19 patients. Finally, we expanded our capacity to 45 beds for patients with COVID-19. The Infectious Disease Ward in our hospital is mainly staffed by physicians on a weekly rotation and nurses on a one- or two-month rotation. In addition, we have provided specific medical care for outpatients with fevers since March 2020. We have implemented basic hospital infection control measures to prevent the spread of COVID-19 according to the manuals (in Japanese) produced by the Japanese Society for Infection Prevention and Control ^3^ and the National Center for Global Health and Medicine ^4^. In brief, we have implemented standard precautions for general patients and have used personal protective equipment (PPE), including N95 masks, face shields, caps, gowns and double gloves, when treating patients with suspected or confirmed COVID-19. We occasionally had a shortage of PPE, which we addressed by using alternative PPE ^5^. Administrative staff also helped medical staff and used the PPE described above when they came into contact with patients with suspected COVID-19.

To evaluate the effectiveness of our hospital’s current countermeasures against nosocomial infections in the context of the COVID-19 pandemic, we investigated the subclinical SARS-CoV-2 infection rate in staff at our hospital by measuring anti-SARS-CoV-2 IgG and identified the risk factors for infection at our hospital, which is accepting COVID-19 patients during the pandemic in Japan.

## Methods

### Study participants, sample size and setting

This was a cross-sectional study to examine the prevalence of anti-SARS-CoV-2 immunoglobulin (Ig) G. The study subjects consisted of 1133 hospital staff in 810 full-time jobs and 323 part-time jobs at Toyonaka Municipal Hospital. They underwent an annual regular health check-up in Japan from June 12 to 19, 2020. We investigated the prevalence of anti-SARS-CoV-2 IgG and the risk factors for seropositivity in those who had and did not have direct contact with patients with confirmed or suspected cases of COVID-19.

We invited all our hospital staff planning to undergo a regular health check-up to participate in this study via the intranet at our hospital. The following individuals were excluded from the study: those who refused to take part in this study; those who did not have enough extra blood drawn to undergo antibody testing; and those who were not identified because they mistyped their personal ten-digit identification code. Finally, in 925 participants, we measured antibodies in extra serum from blood samples taken during the regular health check-up.

To investigate the risk factors for seropositivity, we asked participants to answer an anonymous questionnaire consisting of 14 questions about their background, their involvement with general patients, their involvement with patients with suspected or confirmed cases of COVID-19, and their general condition via the Web using a Google form. The details of this questionnaire are shown in Table 1.

**Table 1.**
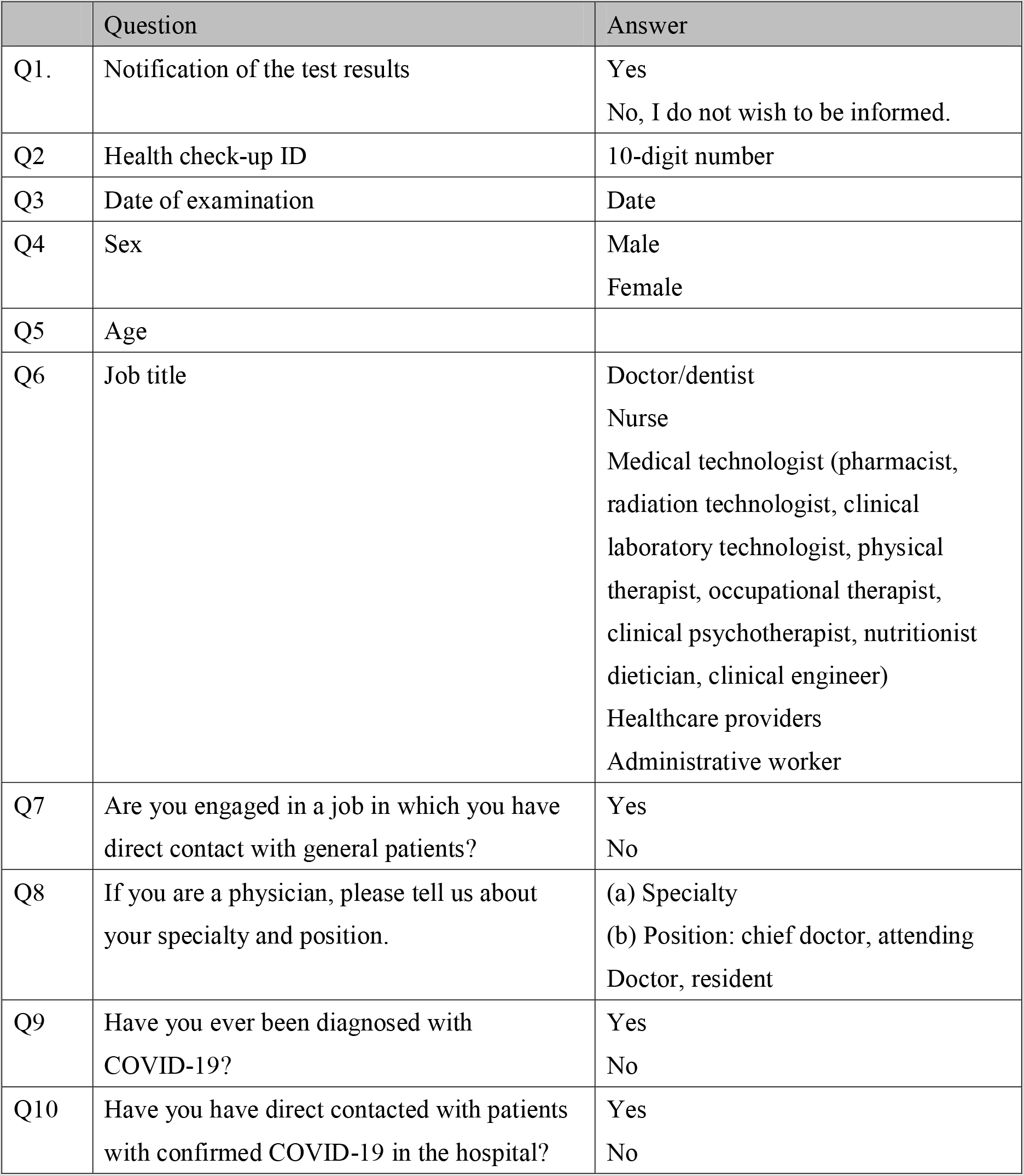
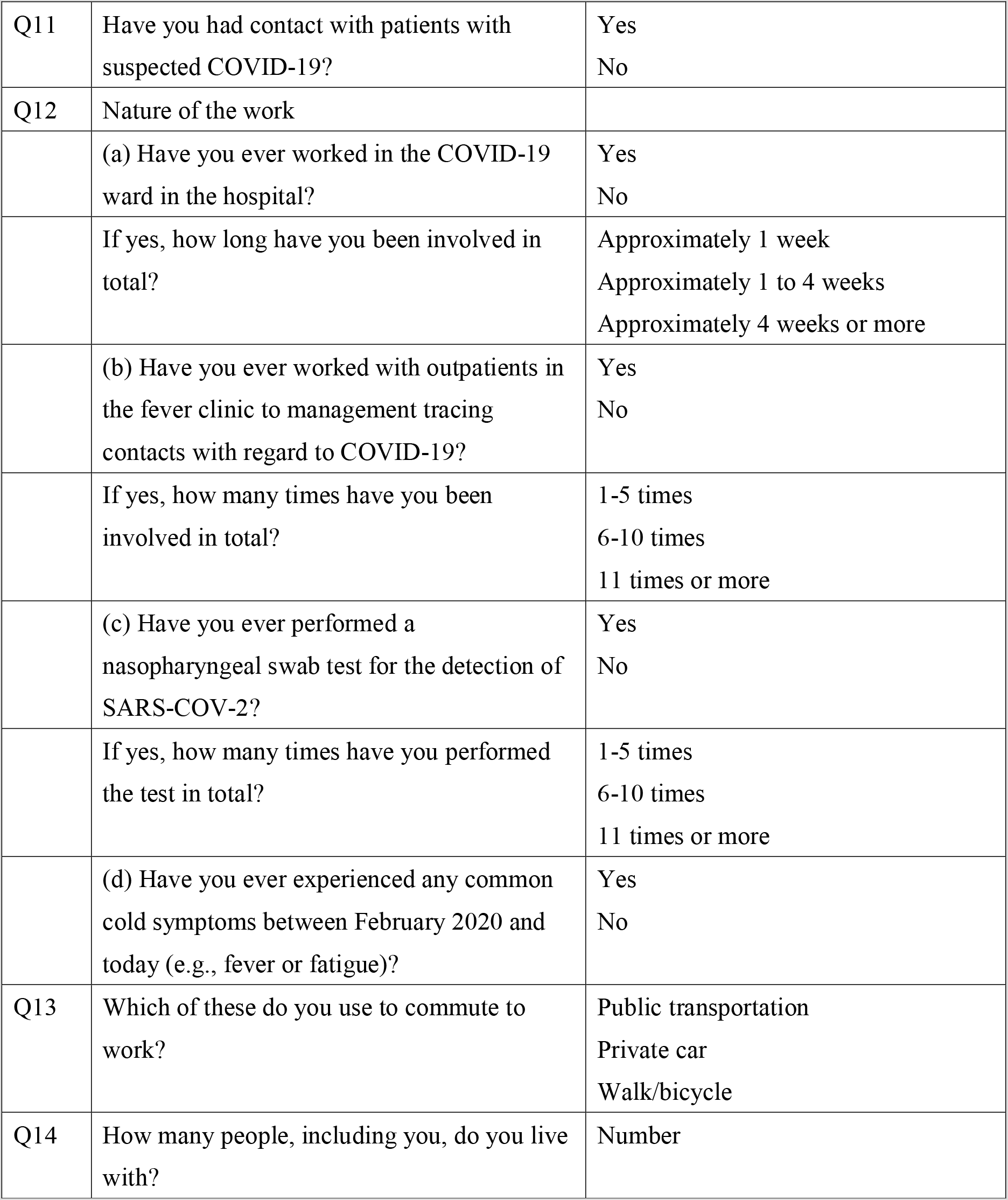
Questionnaire collecting information on the participants’ background, involvement with general patients, involvement with patients with suspected or confirmed cases of COVID-19, and general condition from February 2020 in a multiple-choice format.

|  | Question | Answer |
| --- | --- | --- |
| Q1. | Notification of the test results | Yes<br>No, I do not wish to be informed. |
| Q2 | Health check-up ID | 10-digit number |
| Q3 | Date of examination | Date |
| Q4 | Sex | Male<br>Female |
| Q5 | Age |  |
| Q6 | Job title | Doctor/dentist<br>Nurse<br>Medical technologist (pharmacist, radiation technologist, clinical laboratory technologist, physical therapist, occupational therapist, clinical psychotherapist, nutritionist dietician, clinical engineer)<br>Healthcare providers<br>Administrative worker |
| Q7 | Are you engaged in a job in which you have direct contact with general patients? | Yes<br>No |
| Q8 | If you are a physician, please tell us about your specialty and position. | (a) Specialty<br>(b) Position: chief doctor, attending Doctor, resident |
| Q9 | Have you ever been diagnosed with COVID-19? | Yes<br>No |
| Q10 | Have you have direct contacted with patients with confirmed COVID-19 in the hospital? | Yes<br>No |
| Q11 | Have you had contact with patients with suspected COVID-19? | Yes<br>No |
| Q12 | Nature of the work |  |
|  | (a) Have you ever worked in the COVID-19 ward in the hospital? | Yes<br>No |
|  | If yes, how long have you been involved in total? | Approximately 1 week<br>Approximately 1 to 4 weeks<br>Approximately 4 weeks or more |
|  | (b) Have you ever worked with outpatients in the fever clinic to management tracing contacts with regard to COVID-19? | Yes<br>No |
|  | If yes, how many times have you been involved in total? | 1-5 times<br>6-10 times<br>11 times or more |
|  | (c) Have you ever performed a nasopharyngeal swab test for the detection of SARS-COV-2? | Yes<br>No |
|  | If yes, how many times have you performed the test in total? | 1-5 times<br>6-10 times<br>11 times or more |
|  | (d) Have you ever experienced any common cold symptoms between February 2020 and today (e.g., fever or fatigue)? | Yes<br>No |
| Q13 | Which of these do you use to commute to work? | Public transportation<br>Private car<br>Walk/bicycle |
| Q14 | How many people, including you, do you live with? | Number |

The present study was conducted in accordance with the principles of the Declaration of Helsinki, and approval was obtained from the Institutional Review Board of Toyonaka Municipal Hospital (No. 2020–05–08). We obtained written informed consent from participants prior to the study.

### Sample

All samples were collected and stored at –20°C until use. IgG antibodies against SARS-CoV-2 were detected using a laboratory-based quantitative assay (Abbott ARCHITECT^○R^ SARS-CoV-2 IgG Assay; chemiluminescence microparticle immunoassay; sensitivity: 100%, specificity: 99.6%; Abbott Laboratories, IL, USA) performed on the Abbott Architect i4000SR (Abbott Diagnostics, IL, USA) at the Division of Clinical Laboratory in our hospital according to the manufacturer’s instructions. The Food and Drug Administration (FDA) has not fully authorized any COVID-19 test, but this kit has been authorized for emergency use ^6^.

### Outcomes

The primary outcome was the rate of seropositivity for anti-SARS-CoV-2 IgG. The key secondary outcomes were the rate of seropositivity for anti-SARS-CoV-2 IgG stratified by job title, work tasks, direct contact with general patients, direct contact with patients with suspected or confirmed cases of COVID-19, a history of cold-like symptoms from February to June 2020, commuting methods, and number of cohabitants.

### Statistical analysis

The means ± standard deviations are reported for continuous variables. Categorical variables are summarized as frequencies (percentages) with 95% confidence intervals (CIs) for the rate of seropositivity. A t-test was used to compare age. Differences were assessed by Fisher’s exact test or the chi-square test. All reported P values were two-sided, and P< 0.05 was considered significant. The statistical analyses were performed with JMP statistical software (ver. 14.3, SAS Institute, Inc., Cary, NC, USA).

## Results

Of the 1133 hospital staff who had planned to undergo an annual health check-up at our hospital in June 2020, 926 agreed to participate in the present study. Finally, 925 (81.6%) were tested for anti-SARS-CoV-2 IgG. One person was not tested due to an inadequate amount of serum. There was a female predominance (80.0%). The mean age was 40.0±11.8 years. There were 149 medical doctors or dentists (16.0%), 489 nurses (52.9%), 140 medical technologists (14.2%), 49 healthcare providers (5.3%), and 98 administrative staff members (10.5%) (Table 2).

**Table 2.** Seropositive rate according to characteristics.

|  | N (%) | Seropositive<br>N (%) | 95% CI<br>(%) | P value |
| --- | --- | --- | --- | --- |
| Total number | 925 | 4 (0.43) | 0.17-1.1 |  |
| Sex |  |  |  | 0.1805 |
| Male | 185 (20) | 2 (1.1) | 0.30-3.9 |  |
| Female | 740 (80) | 2 (0.27) | 0.074-1.0 |  |
| Age group, years |  |  |  | 0.5232 |
| 20-29 | 234 (25) | 0 |  |  |
| 30-39 | 225 (24) | 1 (0.44) | 0.079-2.5 |  |
| 40-49 | 250 (27) | 1 (0.40) | 0.071-2.2 |  |
| 50-69 | 216 (25) | 2 (0.93) | 0.25-3.3 |  |
| Job title |  |  |  | 0.2809 |
| Doctor or dentist | 149 | 2 (1.3) | 0.37-4.8 |  |
| Nurse | 489 | 1 (0.20) | 0.036-1.1 |  |
| Medical technologist | 140 | 0 (0) |  |  |
| Healthcare provider | 49 | 0 (0) |  |  |
| Administrative staff | 98 | 1 (1.0) | 0.18-5.6 |  |
| Doctor specialty* |  |  |  | 1.000 |
| Internist specialty | 59 | 1 (1.7) | 0.30-9.0 |  |
| Surgical specialty | 85 | 1 (1.2) | 0.21-6.4 |  |
| Doctor position |  |  |  | 0.2467 |
| Chief doctor | 40 | 0 (0) |  |  |
| Attending doctor | 63 | 2 (3.2) | 0.88-11 |  |
| Resident | 46 | 0 (0) |  |  |

CI; confidential interval \*Missing data N=5

Overall, 4 participants were positive for anti-SARS-CoV-2 IgG (0.43%, 95% confidential interval (CI); 0.17–1.1%). Seropositive participants were significantly older than seronegative participants (52.8±6.8 vs. 40.0±11.8, P=0.0309), but sex was not significantly different (males: 50% (2/4) vs. 19.9% (183/921), P=0.1805). Table 2 shows the prevalence of seropositivity for anti-SARS-CoV-2 IgG stratified by the participants’ characteristics. Doctors and dentists had a slightly higher rate of seropositivity (1.3%) than people in other jobs, but there was no significant difference in rates among people with different job titles. No hospital staff responded that they had been diagnosed with COVID-19 since February 2020.

Table 3 shows the prevalence of seropositivity for anti-SARS-CoV-2 IgG based on exposure to patients with COVID-19. Subjects who had experienced common cold symptoms from February 2020 to June 2020 had a slightly higher seropositivity rate, but the difference was not significant. There were no significant differences in rates based on any other factors.

**Table 3.** Seropositive rate according to work at the hospital.

| Question | N (%) | Seropositive<br>N (%) | 95% CI<br>(%) | P value |
| --- | --- | --- | --- | --- |
| Total number | 925 | 4 (0.43) | 0.17-1.1 |  |
| Direct contact with patients |  |  |  | 1.0000 |
| Yes | 758 (81.9) | 4 (0.53) | 0.21-1.3 |  |
| No | 167 (18.1) | 0 (0) |  |  |
| Direct contact with confirmed<br>COVID-19 patients |  |  |  | 0.3282 |
| Yes | 267 (28.9) | 2 (0.75) | 0.21-2.7 |  |
| No | 658 (71.1) | 2 (0.30) | 0.083-1.1 |  |
| Direct contact with suspected<br>COVID-19 patients |  |  |  | 1.000 |
| Yes | 452 | 2 (0.44) | 0.12-1.6 |  |
| No | 473 | 2 (0.42) | 0.11-1.5 |  |
| Work experience in the ward for<br>COVID-19 patients? |  |  |  | 1.000 |
| Yes | 216 | 1 (0.46) | 0.082-2.6 |  |
| No | 709 | 3 (0.42) | 0.14-1.2 |  |
| Work experience at fever<br>outpatient clinic related to the<br>management of contact with<br>suspected COVID-19 patients? |  |  |  | 1.000 |
| Yes | 145 | 0 (0) |  |  |
| No | 780 | 4 (0.51) | 0.20-1.3 |  |
| Performance of a nasopharyngeal<br>swab test for *the* detection of<br>SARS-CoV-2 at fever outpatient<br>clinic related to the management<br>of contact with suspected<br>COVID-19 patients? |  |  |  | 1.000 |
| Yes | 72 | 0 (0) |  |  |
| No | 853 | 4 (0.47) | 0.18-1.2 |  |
| Common cold symptoms |  |  |  | 0.0716 |
| Yes | 110 | 2 (1.8) | 0.50-6.4 |  |
| No | 815 | 2 (0.25) | 0.067-0.89 |  |
| Commute method |  |  |  | 0.3682 |
| Public transportation | 396 | 3 (0.76) | 0.26-2.2 |  |
| Private car | 214 | 0 (0) |  |  |
| Walk/bicycle | 315 | 1 (0.32) | 0.056-1.8 |  |
| Cohabitant number |  |  |  | 0.0773 |
| 1 | 220 | 0 (0) |  |  |
| 2 | 204 | 3 (1.5) | 0.50-4.2 |  |
| 3 | 188 | 0 (0) |  |  |
| 4 | 207 | 0 (0) |  |  |
| 5 or more | 106 | 1 (0.94) | 0.17-5.2 |  |

CI; confidence interval

## Discussion

During the three months from February 22 to May 31, our hospital accepted 75 patients with confirmed COVID-19 ^7^. We also performed a total of 415 nasopharyngeal swabs for the detection of SARS-CoV-2 during this period, and 61 were positive (14.7%) (data not shown). We have implemented standard precautions when caring for general patients and have used PPE when caring for patients with suspected or confirmed cases of COVID-19. Under the current circumstances, it is important to investigate the subclinical SARS-CoV-2 infection rate in our staff and to evaluate the effectiveness of our nosocomial infection control measures.

This study showed that the prevalence of seropositivity for SARS-CoV-2 IgG as evaluated with a laboratory-based quantitative test (Abbott immunoassay) was 0.43% in our hospital. Considering that the sensitivity of this assay is 100%, the true-positive rate in our hospital should be less than 0.43%. In addition, 0.43% is similar to or less than the proportion (0.54% (16/2970)) identified by the same assay in the general population in Osaka during the same period (7). The results of this study demonstrated that our nosocomial infection control measures have thus far been successful.

Until now, the percentage of the staff of a general hospital with subclinical SARS-CoV-2 infections has remained unclear. Although it may change according to the epidemic condition in the region in which the hospital is located or the number of COVID-19 patients the hospital accepted, this percentage is an important metric for the evaluation of the effectiveness of the nosocomial infection control measures implemented by the hospital. Our hospital is located in the pandemic region in Japan, however, the situation regarding the pandemic is much more severe in China than in Japan ^8^. The strength of the present study was that we could compare the results with large-scale control data in the same region. Osaka is the second-largest metropolitan region in Japan, with a population of 2.67 million. As of July 21, 2020, there was a total of 2541 confirmed cases COVID-19 and 84 related deaths in the Osaka region. Fortunately, the Ministry of Health, Labour and Welfare of Japan conducted a seroprevalence survey in the general population in three different regions in Japan, including Osaka, during the same period and using the same immunoassay from Abbott Laboratories, which revealed seropositive rates of 0.54% (16/2970) in Osaka, 0.2% (4/1971) in Tokyo, and 0.11% (3/3009) in Miyagi ^9^. Therefore, we can compare this seropositive rate in Osaka with our rate. Compared with the seropositive rate of 0.54% in the general population in Osaka, our results indicate that we have thus far successfully managed to avoid hospital-acquired infections.

The Softbank Group Corp. conducted a serological survey from May 12, 2020, to June 8, 2020, which has been the most extensive survey in Japan, identifying 191 positive results in 44,066 volunteers (0.43%), including 38,216 employees at Softbank Group and their family members. This positive rate was similar to our result. However, they used different antibody kits manufactured by Zhejiang Orient Gene Biotech Co., Ltd. (China) and Innovita (China), which are not endorsed by the WHO. In addition, the Softbank Group survey also consisted of 5850 subjects with ties to a hospital, and 105 were positive, for a seropositive rate of 1.79% ^10^. They concluded that health care providers are generally more likely than the general public to be positive for anti-SARS-CoV-2 antibodies ^10^. However, the positive rate of 0.43% in our hospital was similar to that in the general population in Osaka. These data also suggested that our infection control measures appear to have been more effective against SARS-CoV-2 than those of other hospitals.

We obtained some information about the participants’ backgrounds, their involvement with general patients, their involvement with patients with suspected or confirmed cases of COVID-19, and their general condition. We attempted to investigate risk factors associated with seropositivity, but the low positive rate made this impossible. Future studies with a longer period will be needed.

This study has several limitations. First, we could not survey all the staff in our hospital; thus, the prevalence found in this study may not be exact. However, more than 80% of the hospital staff in our hospital, including staff in all jobs, were involved in the present study. We believe the result obtained from this study is very close to the exact value. Second, there is an issue with serological tests. Serological tests do not detect the virus itself and instead reflect the body’s immune response to infection by the virus. Therefore, false-positive results are possible due to cross-reactivity with pre-existing antibodies and other reasons. The specificity of the immunoassay used in this study is reported to be 99.6%, indicating that there could have been four false-positive cases in every 1000 subjects tested ^11^. Although we should consider this limitation of the serological test, we can assume at the very least that the subclinical SARS-CoV-2 infection rate is less than 0.43% in our hospital.

In conclusion, we found that the subclinical SARS-CoV-2 infection rate in our hospital, which treats COVID-19 patients during the pandemic in Japan, is not higher than that in the general population in the same area during the same period. Timely serological screening of a large cohort is essential for achieving control during the pandemic ^12^. Furthermore, hospital-based antibody screening could also help us evaluate and monitor infection control. A longitudinal survey of serum antibodies would be necessary to clarify whether control measures have been effective.

## Data Availability

The data that support the findings of this study are available on request from the corresponding author. The data are not publicly available due to privacy or ethical restrictions.

## Author Contributors

Conceptualization; Nishida T and Iwahashi H, Data curation; Nishida T, Formal analysis; Nishida T and Iwahashi H, Funding acquisition; Nishida T and Iwahashi H, Investigation; Nishida T, Yamauchi K and Kinoshita N, Methodology; Nishida T and Iwahashi H, Project administration; Nishida T and Iwahashi H, Resources; Yamauchi K and Kinoshita N, Supervision; Iwahashi H, Roles/Writing - original draft writing Nishida T, review & editing; Iwahashi H, Okauchi Y, Suzuki N, Inada M, Abe K.

## Declaration of interests

We have no conflicts of interest to declare.

## Funding

Unrestricted research funds from the Department of Gastroenterology and Department of Internal Medicine, Diabetes Center, Toyonaka Municipal Hospital.

## Acknowledgements

We thank Issuikai and Hokenkagakunishinihon for collecting and storing the serum samples from our staff.

## Notes

### Competing Interest Statement

The authors have declared no competing interest.

### Author Declarations

The present study was conducted in accordance with the principles of the Declaration of Helsinki, and approval was obtained from the Institutional Review Board of Toyonaka Municipal Hospital (No. 2020-05-08). We obtained written informed consent from participants prior to the study.

## References

1. Chu J, Yang N, Wei Y et al. Clinical characteristics of 54 medical staff with COVID-19: A retrospective study in a single center in Wuhan, China. J Med Virol 2020; doi 10.1002/jmv.25793.

2. WHO. Coronavirus disease (COVID-19) Situation Report – 176, https://www.who.int/docs/default-source/coronaviruse/situation-reports/20200714-covid-19-sitrep-176.pdf?sfvrsn=d01ce263_2; 2020 [accessed Aug 7, 2020].

3. Japanese Society for Infection Prevention and Control http://www.kankyokansen.org/modules/news/index.php?content_id=328; 2020 [accessed Aug 7, 2020].

4. National Center for Global Health and Medicine, https://www.ncgm.go.jp/covid19/PDF/20200616.pdf; 2020 [accessed Aug 7,2020].

5. Nishida T, Suzuki N, Ono Y et al. How to make an alternative plastic gown during the personal protective equipment shortage due to the COVID-19 pandemic. Endoscopy 2020; doi 10.1055/a-1197-5949.

6. US Food and Drug Administration. EUA Authorized Serology Test Performance, https://www.fda.gov/medical-devices/emergency-situations-medical-devices/eua-authorized-serology-test-performance; 2020 [accessed July 19, 2020].

7. Higuchi T, Nishida T, Iwahashi H et al. Early Clinical Factors Predicting the Development of Critical Disease in Japanese Patients with COVID-19: A Single-Center Retrospective Observational Study. medRxiv 2020; doi 10.1101/2020.07.29.20159442: 2020.07.29.20159442.

8. Chen Y, Tong X, Wang J et al. High SARS-CoV-2 antibody prevalence among healthcare workers exposed to COVID-19 patients. J Infect 2020; doi 10.1016/j.jinf.2020.05.067.

9. Ministry of Health, Labor and Welfare of Japan https://www.mhlw.go.jp/content/000640287.pdf; 2020 [accessed July 6, 2020].

10. Softbank Group, https://group.softbank/system/files/pdf/antibodytest.pdf; 2020 [accessed July 7].

11. Nakamura A, Sato R, Ando S et al. Seroprevalence of Antibodies to SARS-CoV-2 in Healthcare Workers in Non-epidemic Region: A Hospital Report in Iwate Prefecture, Japan. medRxiv 2020; doi 10.1101/2020.06.15.20132316: 2020.06.15.20132316.

12. Abbasi J The Promise and Peril of Antibody Testing for COVID-19. Jama 2020; doi 10.1001/jama.2020.6170.

